# Seroprevalence of antibodies to SARS-CoV-2 is particularly high in patient population with ocular surface diseases

**DOI:** 10.1101/2020.09.22.20198465

**Authors:** Shengjie Li, Yichao Qiu, Li Tang, Zhujian Wang, Wenjun Cao, Xinghuai Sun

**Affiliations:** Clinical Laboratory, Eye & ENT Hospital, Fudan University, Shanghai, 200031, China; Eye Institute and Department of Ophthalmology, Eye & ENT Hospital, Fudan University, Shanghai, 200031, China

## Abstract

Coronavirus disease 2019 (COVID-19), a novel respiratory disease caused by severe acute respiratory syndrome coronavirus 2 (SARS-CoV-2), emerged in December 2019. It is vitally important to perform a seroprevalence study to estimate the percentage of people with SARS-CoV-2 antibodies in the patient population with ocular surface diseases. IgG and IgM antibodies against the SARS-CoV-2 spike protein and nucleoprotein were measured using a commercially available magnetic chemiluminescence enzyme immunoassay kit. Throat swabs were tested for SARS-CoV-2 RNA in a designated virology laboratory. A total of 6, 414 individuals were enrolled. All participants throat swabs were RT-PCR-negative for SARS-CoV-2. Seroprevalence in the patient population was 0.31%. And the seroprevalevce in ocular surface diseases, no-ocular surface diseases and no-ocular diseases are 1.82% (6/330), 0.22% (10/4614) and 0.27% (4/1470) respectively.

---

Coronavirus disease 2019 (COVID-19), a novel respiratory disease caused by severe acute respiratory syndrome coronavirus 2 (SARS-CoV-2), emerged in December 2019.^1^ It is noteworthy that some patients have ocular manifestations, including conjunctival hyperemia, chemosis, epiphora, and increased secretions.^2^ SARS-CoV-2 receptor angiotensin-converting enzyme 2 (ACE2) and entry protease transmembrane serine protease 2 are strongly detected in human ocular surfaces, indicating that they might provide SARS-CoV-2 with a portal of entry.^3^ Thus, it is vitally important to perform a seroprevalence study to estimate the percentage of people with SARS-CoV-2 antibodies in the patient population with ocular surface diseases.

## Methods

This study included individuals with various ocular diseases in the Eye and ENT Hospital of Fudan University (Fenyang branch and Pujiang branch) from May to October 2020. Ethics approval was obtained from the Medical Ethics Committee of the Eye and ENT Hospital of Fudan University. The study adhered to the principles of the Declaration of Helsinki. Informed consent was obtained from all participants.

IgG and IgM antibodies against the SARS-CoV-2 spike protein and nucleoprotein were measured using a commercially available magnetic chemiluminescence enzyme immunoassay kit (Bioscience, Chongqing, China). Antibody levels were expressed as the ratio of the chemiluminescence signal to the cutoff (S/CO) value. An S/CO value higher than 1 for either IgG or IgM was considered positive. If the S/CO value was higher than 0.7 but lower than 1.1, the serum sample was re-analyzed. Serological assays for SARS-CoV-2 infection have been validated in a previous study.^4^ Throat swabs were tested for SARS-CoV-2 RNA in a designated virology laboratory (KingMed Diagnostics, Shanghai, China) using RT-PCR assays.

## Results

A total of 6, 414 individuals were enrolled. All participants’ throat swabs were RT-PCR-negative for SARS-CoV-2. The levels of IgM and IgG antibodies are presented in Figure 1.

**Figure 1.**
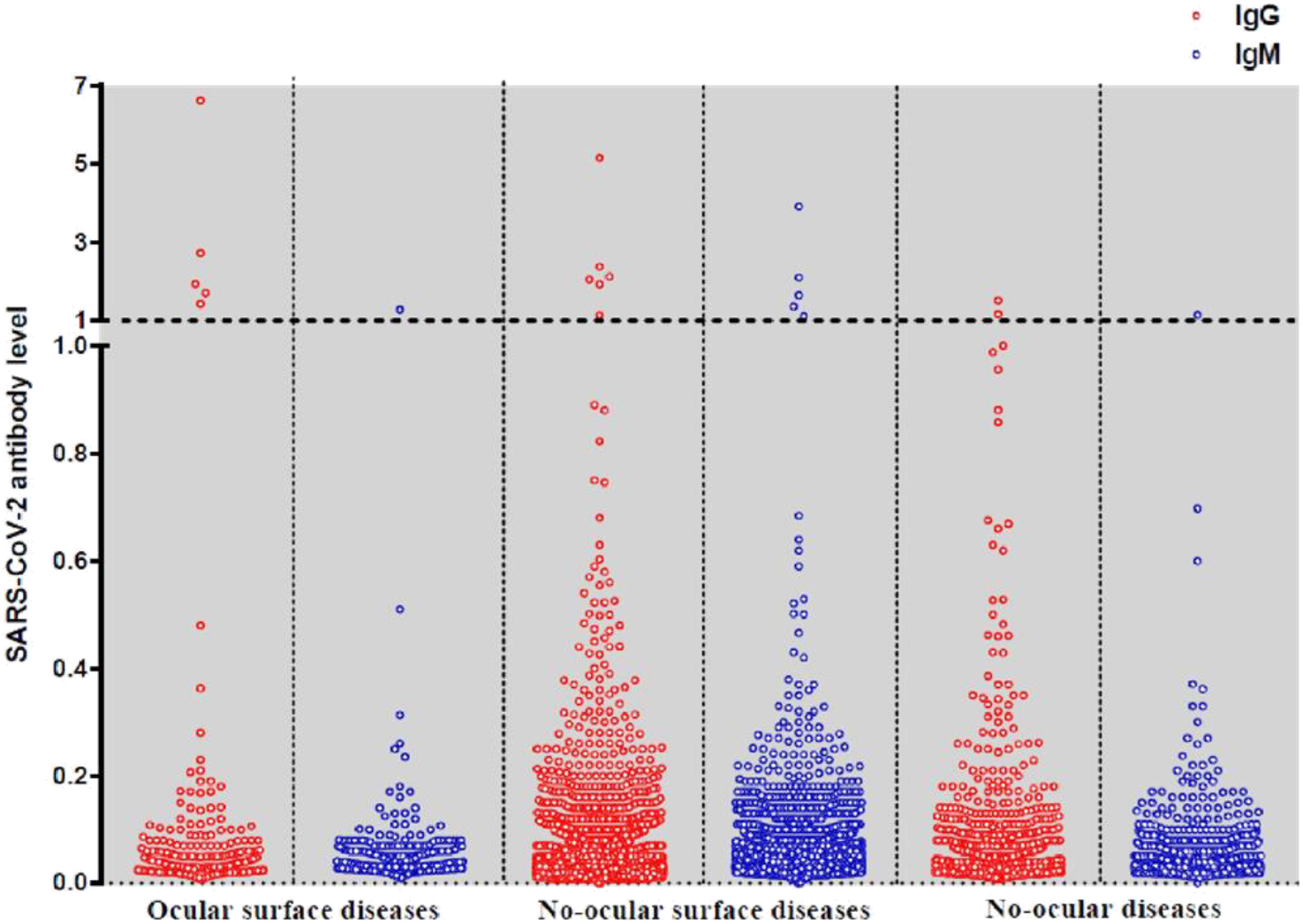
Levels of IgG and IgM antibodies against the nucleoprotein and a peptide from the spike protein of SARS-CoV-2. Each dot represents the level of IgG (red) or IgM (blue) antibody for an individual. The antibody level is expressed as a ratio calculated by dividing the chemiluminescence signal by the cutoff value (S/CO). The dotted line means that the antibody level is at the cutoff point (S/CO = 1).

Seroprevalence in the patient population was 0.31%. And the seroprevalevce in ocular surface diseases, no-ocular surface diseases and no-ocular diseases are 1.82% (6/330), 0.22% (10/4614) and 0.27% (4/1470) respectively (Table 1).

**Table 1.** Positive rates of the serological tests in patients having ocular disease in different part of ocular region and having no ocular disease

|  | n | IgG<sup>+</sup> | IgM<sup>+</sup> | IgM<sup>+</sup> or IgG<sup>+</sup> |
| --- | --- | --- | --- | --- |
| Ocular surface diseases | 330 | 1 (0.30%) | 5 (1.52%) | 6 (1.82%) |
| No-ocular surface diseases | 4614 | 5 (0.12%) | 6 (0.13%) | 10 (0.22%) |
| No ocular disease | 1470 | 1 (0.07%) | 3 (0.20%) | 4 (0.27%) |
| Total | 6414 | 7 (0.12%) | 14 (0.22%) | 20 (0.31%) |

In the 20 seroprevalence positive patients,there are 15 females and 5 males. Patients with IgM seropositivity were twice as many as those with IgG seropositivity. Only one patient had both IgG and IgM seropositivity.

## Discussion

Our results showed that seroprevalence in ocular surface diseases group were particularly higher than other groups. Clinical and scientific evidence suggests that the human ocular surface is susceptible to SARS-CoV-2 infection.^5^ Our previous study suggested that SARS-CoV-2 receptor ACE2 is expressed in human, especiallydiseased conjunctival tissue.^6^ Therefore, it is possible that in patients with ocular diseases— especially ocular surface diseases, such as keratitis and conjunctival cyst— ACE2 is upregulated, making them more susceptible to SARS-CoV-2 infection.^6^ This may be the reason that seroprevalence was particularly high in patient population with ocular surface diseases.

Our study has certain limitations. It was a single-center study, and the patient sample was not large enough, which might have influenced the seroprevalence estimation. Moreover, if samples were collected before infected individuals had a serological response, the serological assays may have produced false negatives. In conclusion, seroprevalence was particularly high in patients with ocular surface diseases. Therefore, increasing awareness of eye protection during the pandemic is necessary, especially for individuals with ocular surface diseases.

## Data Availability

The raw data supporting the conclusions of this article will be made available by the authors

## Conflict of Interest Disclosures

None reported.

## Author Contributions

L SJ, XH S, and C WJ contributed to the study design, data analysis and interpretation, writing of the manuscript and critical revision of the manuscript. Q YC contributed to the writing of the manuscript and critical revision of the manuscript. L SJ contributed to data analysis, interpretation and writing of the manuscript. T L and W ZJ contributed to sample measurements. T L, L SJ and W ZJ contributed to data analysis and validation of serological tests. L SJ and C WJ contributed to clinical management, patient recruitment and data. All authors reviewed and approved the final version of the manuscript.

## Financial support

This work was supported by the Shanghai Sailing Program (18YF1403500), the Shanghai Municipal Commission of Health and Family Planning (20174Y0169 and 201840050), the State Key Program of National Natural Science Foundation of China (81430007), the subject of major projects of National Natural Science Foundation of China (81790641), the Shanghai Committee of Science and Technology of China (17410712500), and the top priority of clinical medicine center of Shanghai (2017ZZ01020) and Shanghai Science and Technology Committee Foundation grant (19411964600).

## Notes

### Competing Interest Statement

The authors have declared no competing interest.

### Author Declarations

The Medical Ethics Committees of Eye and ENT Hospital of Fudan University approved this study

